# Genetic Predisposition to High Blood Pressure and Out-of-Office Hypertension: Insights from a Population Sample in Liechtenstein

**DOI:** 10.1101/2022.12.21.22282423

**Authors:** Sukrit Narula, Pedrum Mohammadi-Shemirani, Stefanie Aeschbacher, Michael R. Chong, Ann Le, Sébastien Thériault, Kirsten Grossman, Guillaume Paré, Lorenz Risch, Martin Risch, David Conen

**Affiliations:** Department of Internal Medicine, Yale University, New Haven, Connecticut, 06510, USA; Department of Health Research Methods, Evidence, and Impact, McMaster University, Hamilton, Ontario, L8S 4K1, Canada; Population Health Research Institute, Hamilton, Ontario, L8L 2X2, Canada; Thrombosis and Atherosclerosis Research Institute, David Braley Cardiac, Vascular and Stroke Research Institute, Hamilton, Ontario, L8L 2X2, Canada; Department of Medical Sciences, McMaster University, Hamilton, Ontario, L8S 4K1, Canada; Department of Medicine, Division of Cardiology, University Hospital Basel, Basel, Switzerland; Cardiovascular Research Institute Basel, University Hospital Basel, Basel, Switzerland; Department of Molecular Biology, Medical Biochemistry and Pathology, Faculty of Medicine, Université Laval, Québec City, Québec, G1V 0A6, Canada; Faculty of Medical Sciences, Private University Triesen, Liechtenstein; Dr Risch Medical Laboratory, Vaduz, Liechtenstein; Department of Pathology and Molecular Medicine, McMaster University, Hamilton, Ontario, L8S 4K1, Canada; Center of Laboratory Medicine, University Institute of Clinical Chemistry, University of Bern, Bern, Switzerland; Dr Risch Medical Laboratory, Buchs, Switzerland; Central Laboratory, Kantonsspital Graubünden, Chur, Switzerland

**Keywords:** Family Medical History, Ambulatory Blood Pressure Monitoring, Human Genetics, Cardiovascular Disease, Risk Factors

## Abstract

Genetic predisposition is a risk factor for office hypertension. We tested whether genetic background could identify individuals with ambulatory daytime hypertension in a sample of white Europeans from Liechtenstein. We evaluated two measures of predisposition to hypertension: family history and polygenic risk scores (PRS). Our analytic sample contained 1444 participants aged 25 to 41. Of the participants, 12% had office hypertension, while 37% had out-of-office hypertension. The correlation between blood pressure PRS and family history of hypertension was low (R^2^ = 4.96×10^−3^), but both were strongly associated with ambulatory blood pressure (2.2 mmHg per 1 SD increase [95% CI: 1.6, 2.7] & 2.4 mmHg increase with positive family history [95% CI: 1.3, 3.4], respectively). The PRS provides incremental improvement in predicting ambulatory systolic blood pressure beyond a validated blood pressure prediction score (ΔAIC = - 33), whereas family history does not (ΔAIC = 1). However, the difference in performance between a baseline prediction algorithm for identifying ambulatory systolic daytime hypertension (positive likelihood ratio of 6.87 [95% CI: 5.56, 8.49]; negative likelihood ratio of 0.45 [95% CI: 0.39, 0.51]) and the same model with PRS integrated (positive likelihood ratio of 7.69 [95% CI: 6.18, 9.57]; negative likelihood ratio of 0.43 [95% CI: 0.37, 0.49]) was modest. In conclusion, in a white European sample from Liechtenstein, PRS and family history are distinct constructs that are associated with increased clinical and ambulatory blood pressure. Unlike family history, polygenic risk scores provide incremental information in the identification of individuals with ambulatory hypertension. However, these gains are modest and warrant further development to improve predictive utility at the point-of-care.

## Introduction

Organ damage and clinical events resulting from arterial hypertension can be delayed with preemptive intervention, however hypertension remains underdiagnosed especially in young and apparently health populations.^1–4^ Appropriate identification of those with hypertension requires clinicians to be able to identify those who likely are hypertensive outside of the office regardless of in-office hypertensive status. Indeed blood pressure (BP) derived from ambulatory measurements has a stronger association with mortality, long term-clinical events, and prognostic surrogate endpoints than BP derived from clinic visits.^5–7^ Better identifying these patients would help providers prioritize who needs closer blood pressure follow-up and more intense risk factor modification.

Given that barriers still persist for more widespread use of out-of-office BP measurement, ambulatory BP prediction models (such as the PROOF-BP algorithm) have been developed and validated, particularly in middle-aged and older populations with cardiovascular comorbidities.^8,9^ In younger and healthier adults, some of the risk factors used in models such as PROOF-BP have yet to manifest and it is plausible that other risk factors (such as genetic predisposition) may further augment these prediction models to identify those with ambulatory hypertension.

Two methods to ascertain genetic predisposition to elevated BP build off of the heritability of BP: polygenic risk scores (PRS) and family history of hypertension (FHx). PRS is a single number that captures the predisposition of an individual to a trait attributable to genetic variants and instances of its use have been widely studied in chronic disease.^10^ Familial inheritance is a classic risk factor assumed to be a surrogate measure of genetic predisposition to chronic disease despite the fact that those with family history of disease may also be exposed to *environments* that increase susceptibility. A direct comparative analysis of FHx and PRS has yet to be conducted in the context of identifying those with both in-office and out-of-office hypertension. In this study, we sought to characterize two facets of genetic predisposition to elevated BP (BP PRS and self-reported FHx of hypertension) as predictors of office-based and ambulatory blood pressure. We hypothesize that incorporation of these tools can improve detection of those with ambulatory hypertension.

## Methods

### Participants

We performed our analysis on the baseline samples of the genetic and phenotypic determinants of BP and other cardiovascular risk factors (GAPP) study. In short, GAPP is a population-based cohort consisting of residents of the Principality of Liechtenstein with data collection ongoing. The sample is composed of healthy individuals between the ages of 25 and 41 at baseline with no history of major illness, including cardiovascular disease or treated diabetes mellitus. Full description of inclusion and exclusion criteria as well as other methodological details surrounding study recruitment and variable collection can be found in the previously published protocol.^11^ Written informed consent was obtained from each participant and the study was conducted in accordance with a protocol approved by the local ethics committee. A total of 2170 individuals were recruited for the GAPP study between years 2010 and 2013 and 1444 participants were analyzed for this study after removing participants who failed genomic quality control (n=649 [29%] excluded; see quality control procedures below) or had missing covariate traits (n=76 [3.5%]; complete case analysis).

### Procedures

#### BP Ascertainment

Conventional office BP was measured in all participants by a trained study nurse using the Microlife BP3AG1 device with appropriate cuff size after at least 5 minutes of rest for each participant in a quiet setting. Three measurements were obtained in a seated position on the non-dominant arm.

Twenty-four hour ambulatory BP was measured with a validated Schiller BR-102 automated device fitted to the participant’s non-dominant arm.12 Over the designated 24-hour period, BP was measured every 15 minutes during daytime (between the hours of 0730 and 2200) and every 30 minutes during nighttime (between the hours of 2200 and 0730). All participants were instructed to engage in their usual activities during the measurement period, but to keep their arm still during recordings. Ambulatory BP recordings were repeated if <80% of possible recordings were available. Daytime and nighttime measurements were additionally ascertained through a patient diary kept by each participant during the recording period. Individuals were classified according to their out-of-office hypertension status (daytime ambulatory BP ≥ 135/85 mmHg) and in-office-hypertension status (office BP ≥140/90 as determined by the average of the last two office measurements).

#### Sample Collection and Genotyping Pipeline

Venous blood sample collection was performed by a trained study nurse and samples were centrifuged and immediately stored in a specialized freezer (−80° C). Genotyping was done using the HumanCoreExome BeadChip designed by Illumina Inc. (San Diego, CA). Standard genotyping quality control procedures were undertaken. Analysis was limited to unrelated participants with European ancestry (as determined by self-reported ethnicity and genotype-based principal components analysis). Additional individual level checks were performed for sex inconsistencies and outlier heterozygosity rates. Genetic markers with increased missing call rate (>5%), Hardy Weinberg equilibrium departures (p-value < 1*10^−6^), and low minor allele frequency (minor allele frequency< 1%) were excluded.

#### Genetic Predisposition Ascertainment

We evaluated two traits of genetic predisposition to elevated BP: the polygenic risk score using external weights from publicly available GWAS and self-reported FHx of hypertension.

The BP polygenic risk score (PRS) is a function of the GWAS SNP-phenotype effect estimate as well as the genotype of a given individual at that particular locus. These two pieces of information are combined to yield a single scalar value in each participant. This value can be considered representative of the genetic risk captured by genetic variants common in the population (minor allele frequency >0.01). We used PRS from weights available from the large GWAS meta-analysis (n= 757,601) of the UK Biobank study and the International Consortium of Blood Pressure conducted by Evangelou et al available through the PGS catalog (PGS ID: PGS002257 and PGS002257).^13^ We note that these weights/effect parameters were derived from the relationship of each SNP with *office* derived average systolic BP (in the case of the systolic BP PRS) and diastolic BP (in the case of the diastolic BP PRS). After harmonization, our systolic BP PRS used 867 SNPs and our diastolic BP PRS used 868 SNPs.

FHx of hypertension was classified based on participant self-report. A participant was denoted as having a positive FHx if they reported at least one first-degree family member having hypertension.

### Statistical Analysis

Baseline characteristics were presented with median and interquartile range for continuous variables and numbers with proportions for categorical variables. For each analysis involving PRS, we utilized the concordant BP PRS i.e. the PRS derived from the *diastolic* BP GWAS summary statistics when evaluating the ambulatory diastolic BP outcome and a PRS derived from *systolic* BP GWAS summary statistics when evaluating the ambulatory systolic BP outcome.

#### Assessing the Relationship Between PRS/FHx and BP Traits

We examined the relationship of FHx and PRS with BP traits in two models: 1) a minimally adjusted model (age with a cube root transform, sex); 2) a risk factor adjusted model (age with a cube root transform, sex, hemoglobin A1c, low density lipoprotein cholesterol, high density lipoprotein cholesterol, triglycerides, body mass index, smoking status) with mutual adjustment for PRS and FHx. Effect measures for PRS were reported per 1 standard deviation (sample) increase.

#### Assessing PRS and FHx as an Incremental Predictor of Ambulatory Hypertension

We calculated the positive and negative predictive value of both PRS and FHx as it relates to identifying out-of-office systolic and diastolic hypertension. To do this, we evaluated the predictive value of FHx and PRS relative to the PROOF-BP algorithm, a validated clinical model for out-of-office BP that uses as features: demographics, clinical risk factors, and three office BP readings.^8,14,15^. In other words, we compared the three models: 1) PROOF-BP, 2) PROOF-BP + PRS, and 3) PROOF-BP + FHx using the Akaike Information Criteria (AIC) and root mean square error (RMSE) as metrics of model performance. We also calculated positive (sensitivity/[1-specificity]) and negative ([1-sensitivity]/specificity) likelihood ratios for ambulatory systolic and diastolic hypertension classification using the model outputs.

Analyses were performed using the statistical software R version 4.0.2 (Vienna, Austria).

## Results

Participant characteristics are presented in Table 1. The median age in our study was 37 years (IQR: 31, 40). 588 participants (41%) self-reported a FHx of hypertension in a first degree relative, 177 showed either office systolic or diastolic hypertension (12%), and 530 showed either ambulatory systolic or diastolic hypertension (37%). FHx and systolic BP PRS were uncorrelated (R^2^ = 4.96×10^−3^). We observed a similar pattern with FHx and diastolic BP PRS (R^2^ = 5.74×10^−3^).

**Table 1:**
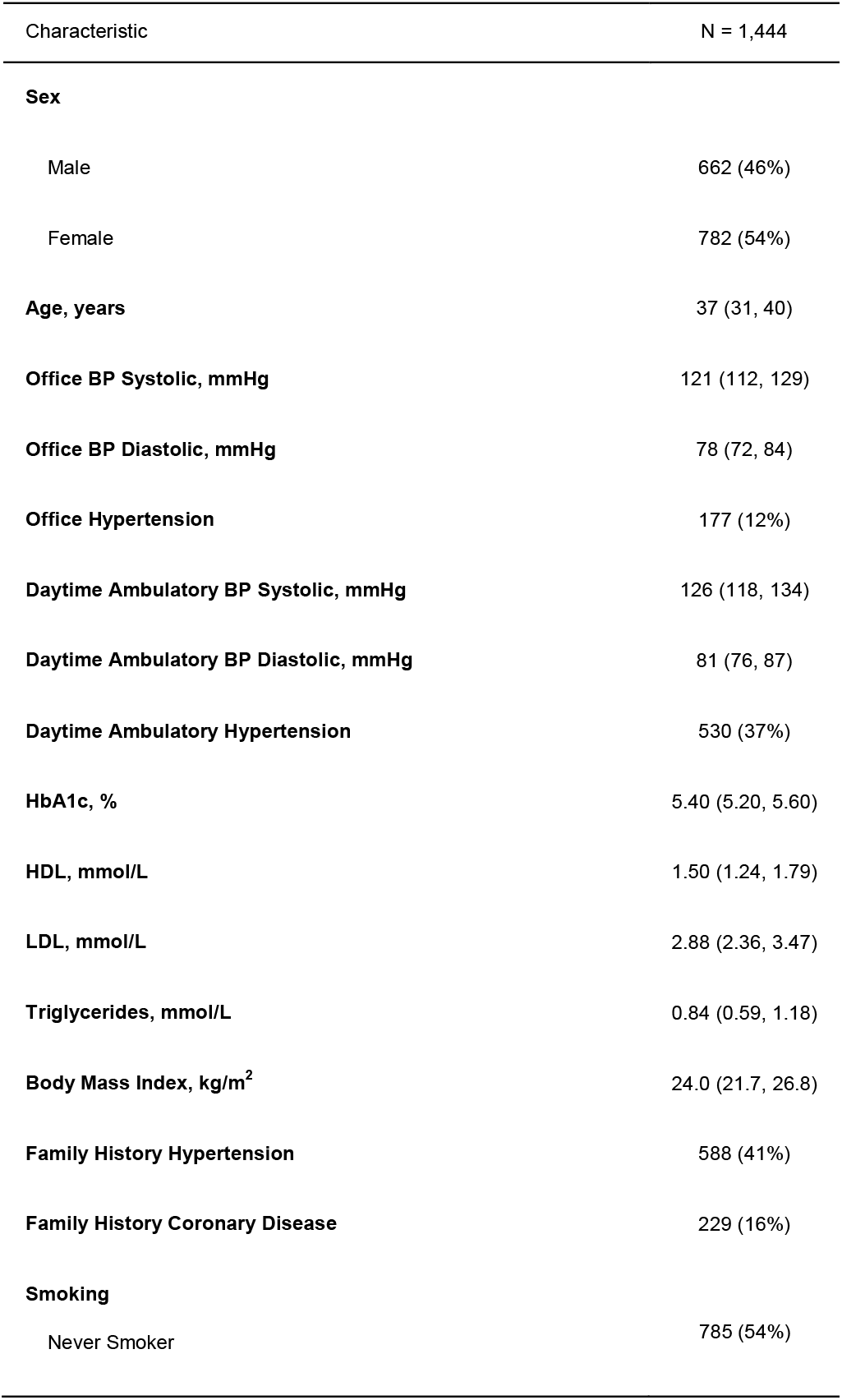
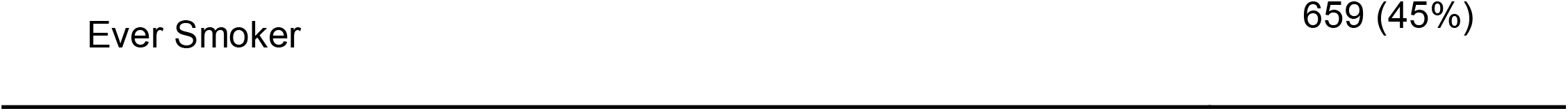
Participant Characteristics

The relationships between genetic predisposition traits and BP are listed in Table 2. PRS was associated with an increased office-based systolic BP (1.8 mmHg per 1 SD increase; 95% CI: 1.3, 2.4) and diastolic BP (1.5 mmHg per 1 SD increase; 95% CI: 1.1, 1.9). PRS was also associated with a higher ambulatory daytime systolic BP (2.2 mmHg per 1 SD increase; 95% CI: 1.6-2.7) and diastolic BP (1.6 mmHg per 1 SD increase; 95% CI: 1.2-2.0). Positive FHx was associated with a 3.5 mmHg higher office systolic BP (95% CI: 2.4-4.7), a 2.6 mmHg higher office diastolic BP (95% CI: 1.8-3.5), a 2.4 mmHg higher ambulatory daytime systolic BP (95% CI: 1.3-3.4), and a 1.8 mmHg higher ambulatory diastolic pressure (95% CI: 1.0-2.7). We found consistent effects after mutual adjustment for FHx status and PRS in addition to adjustment for other risk factors (Supplementary Table 2). Compared to family history, we found that PRS explains more variation in each BP trait than FHx (Supplementary Table 3).

In our cohort, we found that the uncalibrated PROOF-BP algorithm performs comparably to a naïve model consisting only of average office BP measurements in predicting ambulatory systolic BP, and markedly worse in predicting ambulatory diastolic BP (Table 3). As a result, we refit the PROOF-BP equation using the GAPP participants and assessed the incremental value of genetic predisposition compared to a model with these GAPP-recalibrated coefficients. Compared to the recalibrated PROOF-BP model, the model incorporating the genetic score for systolic BP had an improved fit for prediction of ambulatory daytime systolic BP (change in AIC = -32; see Table 3). Relative to the recalibrated PROOF-BP score, a FHx of hypertension provided no incremental value in predicting ambulatory systolic BP. Compared to the recalibrated PROOF-BP model, the model incorporating the genetic score for diastolic BP had an improved fit for prediction of ambulatory daytime diastolic BP (change in AIC = -22; see Table 3). Likewise, a FHx of hypertension did not improve model fit relative to the recalibrated PROOF-BP for diastolic BP prediction. These findings were also consistent when using average blood pressure as a baseline model rather than the GAPP-recalibrated PROOF-BP model (see Supplementary Table 4). Diagnostic test characteristics for predicting ambulatory hypertension traits are presented in table 4. The recalibrated PROOF BP model had a positive likelihood ratio of 6.87 (95% CI: 5.56, 8.49) and negative likelihood ratio of 0.45 (95% CI: 0.39, 0.51) for identifying individuals with ambulatory systolic hypertension (i.e. >135 mmHg) and a positive likelihood ratio of 4.65 (95% CI: 3.89, 5.56) and negative likelihood ratio of 0.46 (95% CI: 0.41, 0.52) for identifying individuals with ambulatory diastolic hypertension (i.e. >85 mmHg). The PROOF-BP model with PRS integrated had a positive likelihood ratio of 7.69 (95% CI: 6.18, 9.57) and negative likelihood ratio of 0.43 (95% CI: 0.37, 0.49) for identifying individuals with ambulatory systolic hypertension and a positive likelihood ratio of 4.72 (95% CI: 3.94, 5.66) and negative likelihood ratio of 0.46 (95% CI: 0.41, 0.52) for identifying individuals with ambulatory diastolic hypertension.

**Table 2:** Association Between Genetic Predisposition to Elevated Blood Pressure and Blood Pressure Traits

| Exposure | Outcome | Systolic Blood Pressure |  |  | Diastolic Blood Pressure |  |  |
| --- | --- | --- | --- | --- | --- | --- | --- |
|  |  | Beta | 95% CI<sup>†</sup> | p-value | Beta | 95% CI<sup>†</sup> | p-value |
| **PRS\*** | Average Office Blood Pressure Reading | 1.8 | 1.3, 2.4 | <0.001 | 1.5 | 1.1, 1.9 | <0.001 |
| **PRS\*** | Ambulatory Daytime Blood Pressure Reading | 2.2 | 1.6, 2.7 | <0.001 | 1.6 | 1.2, 2.0 | <0.001 |
| Exposure | Outcome | Systolic Blood Pressure |  |  | Diastolic Blood Pressure |  |  |
|  |  | Beta | 95% CI<sup>†</sup> | p-value | Beta | 95% CI<sup>†</sup> | p-value |
| **Family History** | Average Office Blood Pressure Reading | 3.5 | 2.4, 4.7 | <0.001 | 2.6 | 1.8, 3.5 | <0.001 |
| **Family History** | Ambulatory Daytime Blood Pressure Reading | 2.4 | 1.3, 3.4 | <0.001 | 1.8 | 1.0, 2.7 | <0.001 |

\*Polygenic Risk Score = PRS. Beta represents per 1 standard deviation increase. Systolic specific PRS effects represented for systolic blood pressure outcome. Diastolic specific PRS effects represented for diastolic blood pressure outcome.
<sup>†</sup>CI = Confidence Interval; All family history and polygenic risk score models presented above adjusted for cube root of age, sex.

**Table 3:** Model Performance with Recalibration and Extension of Proof BP Algorithm

| Model | Ambulatory Systolic Blood Pressure |  |  | Ambulatory Diastolic Blood Pressure |  |  |
| --- | --- | --- | --- | --- | --- | --- |
|  | Adj R2 | AIC | RMSE | Adj R2 | AIC | RMSE |
| Average Office BP | 53.1% | 10161 | 8.14 | 42.1% | 9480 | 6.43 |
| PROOF BP Original | 53.3% | 10153 | 8.12 | 33.8% | 9673 | 6.88 |
| PROOF BP Recalibrated | 57.4% | 10021 | 7.76 | 46.6% | 9362 | 6.18 |
| PROOF BP Recalibrated + Fam Hx | 57.4% | 10022 | 7.76 | 46.6% | 9362 | 6.17 |
| PROOF BP Recalibrated + PRS | 58.4% | 9989 | 7.67 | 47.5% | 9340 | 6.12 |
| PROOF BP Recalibrated + PRS + Fam Hx | 58.4% | 9990 | 7.67 | 47.5% | 9341 | 6.12 |

Average Office BP = average of last two blood pressure readings; Proof BP RC = Proof BP recalibrated; Fam Hx = Family History; PRS = Polygenic risk score with each score fit to its corresponding phenotype i.e. diastolic blood pressure gene score fit with ambulatory diastolic blood pressure phenotype and systolic blood pressure gene score fit with ambulatory systolic blood pressure phenotype; RMSE = Root Mean Squared Error; AIC: Akaike information criterion; Adj R2 = Adjusted R-squared

**Table 4:**
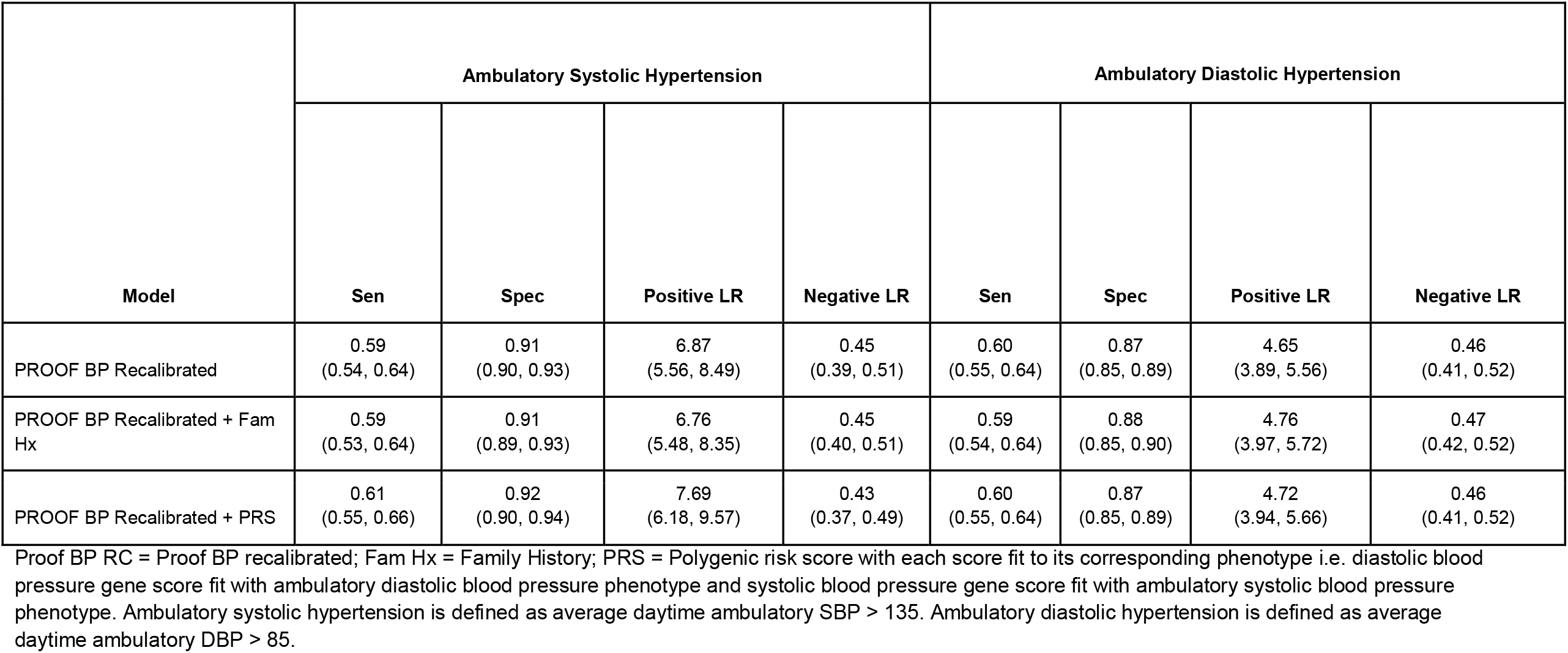
Diagnostic Test Characteristics of Ambulatory Hypertension Recalibrated PROOF-BP and Extensions of PROOF-BP Algorithm

| Model | Ambulatory Systolic Hypertension |  |  |  | Ambulatory Diastolic Hypertension |  |  |  |
| --- | --- | --- | --- | --- | --- | --- | --- | --- |
|  | Sen | Spec | Positive LR | Negative LR | Sen | Spec | Positive LR | Negative LR |
| PROOF BP Recalibrated | 0.59<br>(0.54, 0.64) | 0.91<br>(0.90, 0.93) | 6.87<br>(5.56, 8.49) | 0.45<br>(0.39, 0.51) | 0.60<br>(0.55, 0.64) | 0.87<br>(0.85, 0.89) | 4.65<br>(3.89, 5.56) | 0.46<br>(0.41, 0.52) |
| PROOF BP Recalibrated + Fam Hx | 0.59<br>(0.53, 0.64) | 0.91<br>(0.89, 0.93) | 6.76<br>(5.48, 8.35) | 0.45<br>(0.40, 0.51) | 0.59<br>(0.54, 0.64) | 0.88<br>(0.85, 0.90) | 4.76<br>(3.97, 5.72) | 0.47<br>(0.42, 0.52) |
| PROOF BP Recalibrated + PRS | 0.61<br>(0.55, 0.66) | 0.92<br>(0.90, 0.94) | 7.69<br>(6.18, 9.57) | 0.43<br>(0.37, 0.49) | 0.60<br>(0.55, 0.64) | 0.87<br>(0.85, 0.89) | 4.72<br>(3.94, 5.66) | 0.46<br>(0.41, 0.52) |

Proof BP RC = Proof BP recalibrated; Fam Hx = Family History; PRS = Polygenic risk score with each score fit to its corresponding phenotype i.e. diastolic blood pressure gene score fit with ambulatory diastolic blood pressure phenotype and systolic blood pressure gene score fit with ambulatory systolic blood pressure phenotype. Ambulatory systolic hypertension is defined as average daytime ambulatory SBP > 135. Ambulatory diastolic hypertension is defined as average daytime ambulatory DBP > 85.

## Discussion

In this study, we conducted a thorough evaluation of the relationship between predisposition to elevated BP with both in-office and ambulatory BP traits in a sample with a high burden of ambulatory and masked hypertension. We found a significant relationship between FHx and PRS with elevated BP in both the office and ambulatory setting. We also found that PRS explained more variation in BP phenotypes (both ambulatory and in-office) than FHx. However, for identifying individuals with ambulatory hypertension, PRS provided only modest improvement in prediction, while FHx provided no incremental information. Importantly, FHx had little correlation with PRS suggesting that although both traits are used to capture predisposition to a given trait, they likely provide clinicians with orthogonal information.

An extensive corpus describing the heritable nature of hypertension serves as the basis for understanding the relevance of genetic predisposition in patient assessment.^13,16^ For example, an intergenerational analysis of the Framingham cohort shows that early onset hypertension in antecedent generations is predictive of hypertension in subsequent generations.^17^ Expert statements posit FHx as a risk factor for masked hypertension, along with male sex, diabetes status, and cardiovascular risk factor burden.^18^ However, our findings show that FHx provides no incremental information in the identification of individuals with ambulatory hypertension and are inferior to PRS for identifying those with ambulatory hypertension. Moreover, PRS-based methods have shown promise in the realm of hypertension. An analysis of two Swedish cohorts found that a blood pressure PRS was associated with an increased *incidence* of hypertension with an effect size comparable to that of body-mass index.^19^ Another analysis of the UKBiobank showed that a PRS for office BP was associated with incident cardiovascular disease independent of measured office BP, suggesting that it may provide additional information on cardiovascular risk.^20^ Despite the strong association with blood pressure elevation and its robustness relative to FHx, PRS does not provide enough incremental information, as currently constructed, to improve identification of those with ambulatory hypertensive phenotypes.

Strengths of this study include the large sample size of well-characterized apparently healthy young adults with high participant compliance. This is an otherwise understudied demographic in hypertension cohorts. Previous models developed for ambulatory blood pressure prediction have focused on relatively older patients with higher comorbidity burdens. Most interestingly, we note that the high prevalence of ambulatory hypertension (in particular masked hypertension) in our cohort allowed us to document the informativeness of these ‘predisposition’ traits in those for whom we would otherwise have little clinical suspicion for hypertension. This group would presumably have the largest incremental yield from genetic susceptibility testing. The integration of ambulatory BP monitoring to parse out distinct patterns between office-based and ambulatory measures provides us with unique insights into the relationship between the genetics of BP that have not been described in previous literature. On the other hand, there are limitations that must be pointed out for our work. First, our BP PRS construction is limited to SNPs associated with office BP measurements. GWAS requires large sample sizes for precise estimation of SNP-phenotype relationships, but large sample sizes mean that higher quality phenotypes (like ambulatory BP) are swapped out with noisier phenotypes like office BP. Conduct of large genome wide meta-analyses with higher quality phenotypes like ambulatory BP may improve future gene discovery efforts and the predictiveness of the resulting secondary tools such as PRS. Second, our measure of family history relied on participant self-report. This may be less reliable than previous studies which rely on confirmed inter-generational ascertainment of hypertension status. Despite this, self-report of family history is generally the only information available at the bedside and thus may be more relevant as an operationalization of family history than those with family history observed and verified by study teams. Third, our analysis was limited to PRS constructed from European only source GWAS and applied to a sample of residents from the Principality of Liechtenstein. The consequence of this is that our reported estimates of PRS performance is likely to be optimistic compared to a similar analysis that would be conducted in a more diverse population. Finally, a PRS is not a static entity. In other words, PRS performance for blood pressure can continue to improve with larger GWAS studies, improved phenotyping (as described above), and improved methodology for fitting scores.

### Perspectives

Genetic predisposition traits are strongly associated with BP in both the office and ambulatory setting. PRS provides modest incremental information in ascertaining out-of-office hypertension status in young and healthy individuals, but likely not at a level that sufficiently informs clinical management. FHx, on the other hand, serves as a poor surrogate of genetic predisposition in this setting and does not yield incremental information in identifying those with out-of-office hypertension. This contrasts with recommendations from expert statements suggesting that FHx can be used as a factor in identifying those with masked hypertension. Our investigation suggests PRS may require further progress before it can be used as a tool for identifying ambulatory daytime hypertension. As such, we cannot exclude the possibility of PRS having clinical utility in future applications as methodology improves or GWAS become larger with improved phenotyping.

### Novelty and Significance

#### What is New?

Family history of hypertension (FHx) and polygenic risk score (PRS) for office-based BP were evaluated as predictors for ambulatory daytime BP in a Liechtenstein-based cohort with low burden for hypertension risk factors.

#### What is Relevant?

FHx and PRS are distinct entities with no statistical correlation. Although both traits are associated with increased office and ambulatory daytime BP, PRS is a more robust predictor than FHx. While PRS provided modest incremental value in predicting ambulatory BP, FHx did not improve prediction of ambulatory hypertension status.

#### Summary

PRS is a more informative measure of predisposition to hypertension than family history. The improvement in performance with PRS, however, is modest and warrants further work to increase the utility of PRS at the point-of-care.

## Data Availability

All data produced in the present study are either publicly available, contained in the manuscript, or available upon reasonable request to the authors

## Acknowledgements

We thank those who enrolled in the GAPP study. We also thank the government of the Principality of Liechtenstein, the health ministers, and the Liechtenstein Office of Public Health for their support. Finally, our thanks are especially due to the Princely House of Liechtenstein, which gave decisive support that enabled the initiation of this project.

## Independent data access

The corresponding author and co-authors had full access to the all the data in the study and had final responsibility for the decision to submit for publication

## Non-standard Abbreviations and Acronyms

AIC: Akaike information criterion
BP: blood pressure
FHx: family history
GAPP: Genetic and Phenotypic Determinants of Blood Pressure and Other Cardiovascular Risk Factors
GWAS: genome wide association study
PROOF-BP: PRedicting Out of OFfice Blood Pressure
PRS: polygenic risk score
RMSE: root mean square error

---

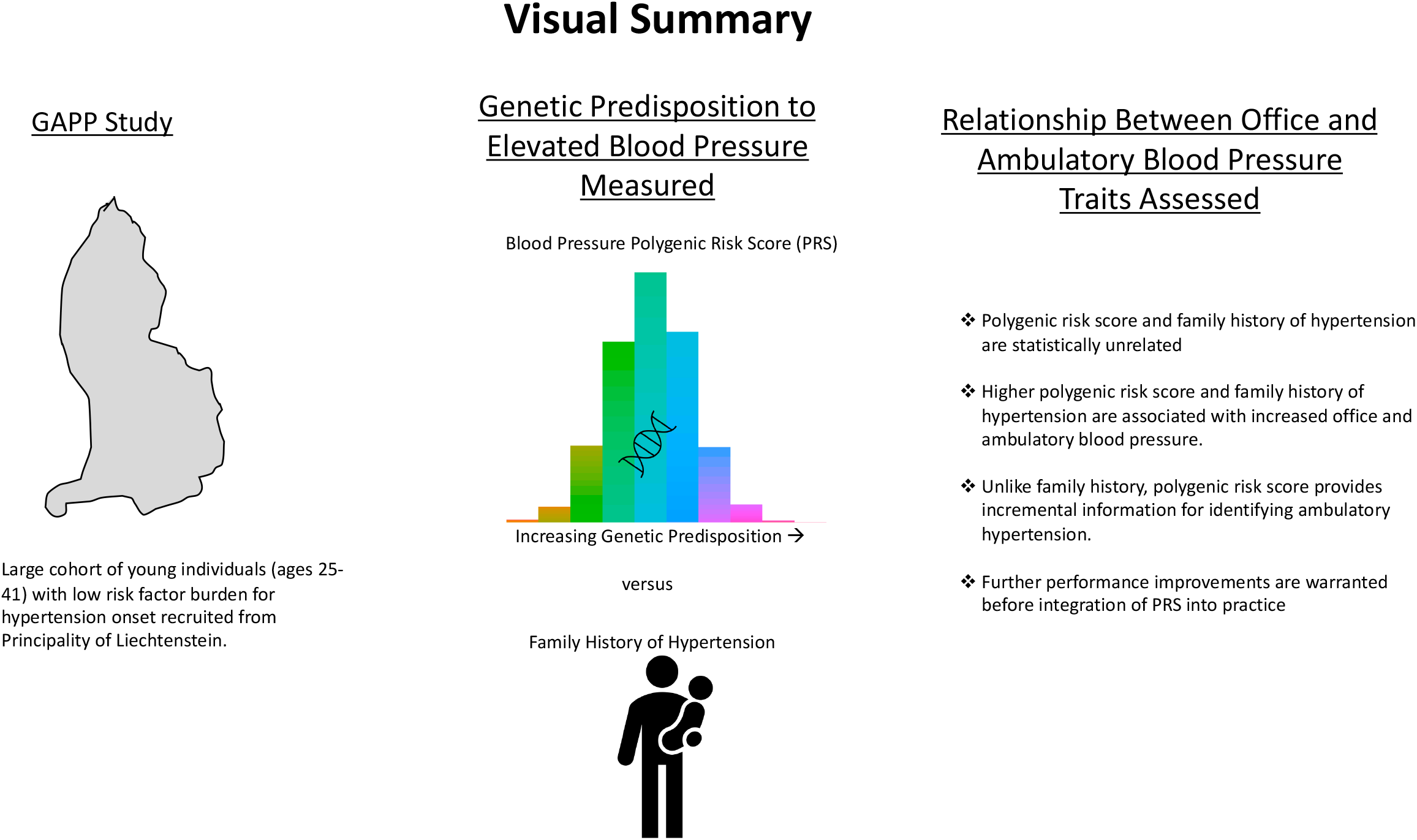

