## Supplementary Appendix for "Genetic Predisposition to High Blood Pressure and Out-of-Office Hypertension: Insights from a Population Sample in Liechtenstein"

**Supplementary Table 1:** Blood Pressure Phenotype Definitions

| **Phenotype** | **Definition** |
| --- | --- |
| Sustained Hypertension | Positive in-office hypertension and positive out-of-office hypertension |
| White Coat Hypertension | Positive in-office hypertension, normotensive out-of-office, without sustained elevation in either systolic or diastolic blood pressure |
| Masked Hypertension | Normotensive in-office, positive out-of-office hypertension, without sustained elevation in either systolic or diastolic blood pressure or white coat effects in either systolic or diastolic blood pressure |
| Normotension | No signs of sustained, white-coat, or masked-hypertension |

Out-of-Office Hypertension = defined as daytime ambulatory blood pressure of greater than or equal to 135 mmHg systolic blood pressure or 85 mmHg diastolic blood pressure; In-office Hypertension = defined as in-office blood pressure greater than or equal to 140 mmHg systolic blood pressure or 90 mmHg diastolic blood pressure for the average of the last two in-office readings

**Supplementary Table 2:** Association Between Genetic Predisposition to Elevated Blood Pressure and Blood Pressure Traits with each model adjusted for hypertension risk factors and mutually adjusted for family history and polygenic risk score

|  |  | **Systolic Blood Pressure** | | | **Diastolic Blood Pressure** | | |
| --- | --- | --- | --- | --- | --- | --- | --- |
| Exposure | Outcome | Beta | 95% CI^†^ | p-value | Beta | 95% CI^†^ | p-value |
| **PRS**^*^ | Average Office Blood Pressure Reading | 1.6 | 1.1, 2.1 | <0.001 | 1.3 | 0.9,1.7 | <0.001 |
| **PRS**^*^ | Ambulatory Daytime Blood Pressure Reading | 2.1 | 1.6, 2.5 | <0.001 | 1.5 | 1.1, 1.9 | <0.001 |
|  |  | **Systolic Blood Pressure** | | | **Diastolic Blood Pressure** | | |
| Exposure | Outcome | Beta | 95% CI^†^ | p-value | Beta | 95% CI^†^ | p-value |
| **Family History** | Average Office Blood Pressure Reading | 2.9 | 1.9, 3.9 | <0.001 | 2.1 | 1.4, 2.9 | <0.001 |
| **Family History** | Ambulatory Daytime Blood Pressure Reading | 1.9 | 0.9, 2.8 | <0.001 | 1.4 | 0.6, 2.1 | <0.001 |

^*^Polygenic Risk Score = PRS. Beta represents per 1 standard deviation increase. Systolic specific PRS effects represented for systolic blood pressure outcome. Diastolic specific PRS effects represented for diastolic blood pressure outcome.

^†^CI = Confidence Interval; All family history and polygenic risk score models presented above adjusted for cube root of age, sex, BMI, smoking status, LDL, Triglycerides, HDL, and were mutually adjusted for PRS and FHx, respectively

**Supplementary Table 3:** Variation Explained (R^2^) by Blood Pressure Genetic Score and Family History with Office/Ambulatory Blood Pressure Traits

|  | Genetic Score | | Family History | |
| --- | --- | --- | --- | --- |
| **Blood Pressure Trait** | **Systolic BP Variation Explained** | **Diastolic BP Variation Explained** | **Systolic BP Variation Explained** | **Diastolic BP Variation Explained** |
| Average Office Blood Pressure Reading | 1.5% | 2.6% | 0.75% | 1.25% |
| Ambulatory Daytime Blood Pressure Reading | 2.6% | 3.2% | 0.18% | 0.76% |

**Supplementary Table 4:** Model Performance Compared to Office Measurements

|  | **Ambulatory Systolic Blood Pressure** | | | **Ambulatory Diastolic Blood Pressure** | | |
| --- | --- | --- | --- | --- | --- | --- |
| **Model** | **Adj R2** | **AIC** | **RMSE** | **Adj R2** | **AIC** | **RMSE** |
| Average Office BP | 53.1% | 10161 | 8.14 | 42.1% | 9480 | 6.43 |
| Average Office BP + PRS | 53.6% | 10147 | 8.10 | 42.6% | 9467 | 6.40 |
| Average Office BP + Fam Hx | 53.1% | 10061 | 8.14 | 42.0% | 9481 | 6.43 |

Average Office BP = average of last two blood pressure readings; Fam Hx = Family History; PRS = Polygenic risk score with each score fit to its corresponding phenotype i.e. diastolic blood pressure gene score fit with ambulatory diastolic blood pressure phenotype and systolic blood pressure gene score fit with ambulatory systolic blood pressure phenotype; RMSE = Root Mean Squared Error; AIC: Akaike information criterion; Adj R2 = Adjusted R-squared
